# Testing the Safety of the Nature Based Microbial Exposure with Atopic Dermatitis Patients – A Randomized, Placebo Controlled, Double-Blinded Pilot Trial

**DOI:** 10.1101/2024.12.27.24319404

**Authors:** Johanna Kalmari, Iida Mäkelä, Laura Kummola, Marja Roslund, Heini Huhtala, Riikka Puhakka, Pekka Autio, Heikki Hyöty, Vesa P Hytönen, Aki Sinkkonen, Olli H Laitinen

**Affiliations:** Faculty of Medicine and Health Technology, Tampere University, Arvo Ylpön katu 34, FI-33520 Tampere, Finland; Uute Scientific Oy, Haartmaninkatu 4, 00290 Helsinki, Finland; Faculty of Biological and Environmental Sciences, University of Helsinki, Niemenkatu 73, FI-15140 Lahti, Finland; Natural Resources Institute Finland Luke, Helsinki and Turku, Finland; Faculty of Social Sciences, Biostatistics Group, Tampere University, Arvo Ylpön katu 34, FI-33520 Tampere, Finland; Aava Medical Center, Annankatu 32, 00100 Helsinki Finland; Fimlab Laboratories, Pirkanmaa Hospital District, Tampere, Finland

**Author notes:** Shared corresponding authorship.

**Keywords:** Atopic dermatitis, microbial exposure, clinical trial, cytokines, antimicrobial peptides

## Abstract

Biodiversity hypothesis posits that it is beneficial for human health to be in contact to microbial biodiversity, and loss of this contact leads to immune-mediated diseases like atopy. Based on this hypothesis, we wanted to study if a lotion containing highly diverse natural microbes is a safe and feasible way to administer nature exposure to humans, and to study the effects of the exposure on the skin. We recruited two groups: 15 people with healthy skin and 12 people with atopic dermatitis. The healthy people were divided to the three groups that used twice daily a lotion containing: 1) 10 % of live microbial extract; 2) 10 % inactivated microbial extract; 3) placebo. The people with atopic dermatitis used a lotion containing 1 % of inactivated microbial extract in one side of their body and a placebo lotion in another side. The daily use of the lotion containing diverse microbial extract was well tolerated by both healthy and atopic people. Microbial extract in lotion did not change the relative abundance of *Staphylococcus* on healthy skin in 14 day-trial and did not change atopic eczema severity, transepidermal water loss or erythema on atopic skin compared to placebo during 28-day trial. Atopic dermatitis patients can benefit from the overall effect of moisturizing lotion and the microbial extract, which together down-regulated pro-inflammatory IL-1β, IL-22, IL-33 cytokines, and antimicrobial peptide hBD-2 on atopic lesions indicating enhanced tolerance and mitigation of eczema. Nature exposure by microbial extract in lotion could complement current atopic dermatitis treatments.

## Introduction

During the last decades exposome, which means all biological, physical, chemical and socio-psychological factors that we are exposed to, has been identified as an important effector to our health ^1^. Exposomic factors, or conversely their absence, are partially responsible for the many diseases including immune-mediated diseases, e.g. autoimmune diseases, asthma, allergies and atopy ^2,3,4^. A starting point for the current study is the biodiversity hypothesis; contacts with microbially rich nature enriches the human microbiome, promotes immune regulation, and protects from immune-mediated diseases ^5–14^. Urbanization, changes in human living environment and lifestyle have decreased our contacts to natural environments, and consequently the lack of exposure to diverse microbial communities can lead to immune system malfunction ^11,15–17^.

The microbiota on the skin and gut is in a dynamic interaction with the environment ^16^. Atopic individuals have been found to have lower environmental biodiversity in the surroundings of their homes and lower diversity of the gammaproteobacteria on their skin compared to healthy individuals ^7^. Cohort studies have also found a correlation between early-life microbiota and atopy ^18^. We have previously gained positive results in biodiversity exposure trials with healthy adults and daycare children, in which contacts to microbiologically rich natural materials have induced beneficial microbiological and immunological outcomes ^12,13,19–21^. A forest based soil and plant material has been shown to diversify the children’s skin and gut microbiome, reduce pathogens, increase the number of regulatory T-cells (Tregs) and change the IL-10/IL-17 cytokine ratio to a more favorable direction ^12,13,20,21^.

Microbial structures are recognized for example by Toll-like receptors (TLR), one class of Pattern Recognition Receptors (PRRs), in epithelial and antigen precenting cells ^22^. The PRRs recognize molecular patterns like microbial proteins and lipopolysaccharides (LPS), and therefore also heat-killed microbes elicit immune responses ^23–26^. It has been demonstrated that high exposure to LPS (endotoxins) in the household is protective against the development of atopy with certain genotype ^27^, and they may also protect from allergies ^28,29^. High overall microbial cell content of drinking water is also associated with lower risk for atopy ^25^. In the absence of microbial stimuli, Tregs, able to attenuate inflammation, and regulatory cytokines are not adequately induced and are likely to disappear, and inflammatory milieu enriching bacteria tolerating inflammatory mediators is generated ^11,30^.

Atopic dermatitis (AD) is a common inflammatory skin disease with a prevalence in adults ranging from 1.2% in Asia to 17.1% in Europe ^31^. Comparison of two populations living at adjacent, socio-economically distinct but geoclimatically similar areas, Finnish and Russian Karelia, has revealed the association of urbanization and western lifestyle to a higher prevalence of atopy, allergy and asthma ^6,32,33,25^. The clinical features of AD include skin inflammation and barrier disruption, which are often associated with abnormal microbial colonization like *Staphylococcus aureus* ^34^. AD skin displays increased expression of many pro-inflammatory cytokines compared to healthy skin like IL-1β, IL-6, IL-8, IL-18 and TNFα involved in innate immunity and IL-4, IL-13, TSLP involved in Th2 signaling ^35,36^. Especially, IL-4 and IL-13 drive barrier dysfunction and keratinocytes in disrupted epidermis produce large amounts of TSLP, IL-17E ja IL-33 conducing to type 2 immune deviation. Type 2 T-cells produce IL-31 which is pruritogen, that increases atopic itch ^34^.

The key part of AD treatment is emollients. The following traditional medical treatments are topical corticosteroids and topical calcineurin inhibitors to reduce inflammation, phototherapy being also one supportive alternative. Biological drugs that target specific inflammatory mediators like IL-4 and IL-13 are considered if atopic dermatitis is severe. ^37^ Some problems are related to medical treatments including high cost and lack of long-term experience for biological drugs ^38^ and skin thinning and decreased effectiveness reported by patients from long-term use of topical corticosteroids ^39^. AD patients prefer to start the treatments with non-medical alternatives, and they are concerned about the adverse effects of medical treatments. Several studies have reported a tendency among patients to minimize the time they use topical corticosteroids. Patients prefer more natural, nonprescription treatments and move on to medical treatments only when AD is more severe. ^39^

As there is a clear need for new treatments we performed a placebo-controlled study to address this need based on the effects of biodiversity exposure in trials with healthy participants with promising immunological outcomes ^12,13,20,21^. The current work aims to study if similar exposure is feasible and safe and could drive beneficial skin-associated immunological changes in participants with AD.

The study was divided into two parts: pretrial with healthy and AD-trial with participants having AD. Both trials were double blinded, placebo controlled and randomized. We found that microbial extract in lotion is safe way to bring nature exposure to healthy and atopic skin. We also noticed that inactivated microbial extract together with moisturizer down-regulate pro-inflammatory cytokines such as IL-1b, IL-22 and IL-33 and antimicrobial peptide human β-defensin 2 on atopic lesional skin. These findings pave way for the utilization of microbe extracts for controlling symptoms associated with sparse contact with nature.

## Material and Methods

### Pretrial: Experimental design

15 healthy volunteers aged between 20–64 participated in the study. Participants were first matched in groups of three according to gender and age. Within each group of three, volunteers were then randomly assigned to two intervention groups and to the control group. One intervention group (n=4) received lotion (Aqualan L, Orion Pharma, Finland) with 10% soil and plant-based material including live microbes and the other group (n=5) with 10% autoclaved soil and plant-based material. The placebo arm (n=6) received lotion mixed with vegetable carbon E153 powder to get the same color like in the other lotions. The soil- and plant-based material has been described in detail in our previous studies ^12,19,40^. The randomization was done by an independent researcher at the University of Helsinki using a random number table. The trial was conducted as double-blinded setup.

The exclusion criteria of the study included: immune deficiencies, immunosuppressive medications, at least three infections within a year that resulted in hospitalization, a disease affecting immune response (e.g., colitis ulcerosa, rheumatoid arthritis, Crohn’s disease), a doctor-diagnosed memory disorder, acute or earlier psychosis or acute depression, cancer diagnosis within the last two years or on-going cancer treatment, diabetes, rash or ulcers in hands, sensitivity to cosmetics or perfumes, age under 18, daily smoking and incompetency. Before the trial, protection against *Clostridium tetani* and immunological health status was confirmed analyzing differential and complete blood count and serum *C. tetani* tetanus toxoid antibodies in a certified hospital laboratory (Fimlab Laboratories, Tampere, Finland). Before starting the trial, a study nurse checked that the skin on participants’ hands were in good condition with no wounds or eczema.

In the pretrial, each group spread lotion on their arm twice a day (before breakfast and bedtime) for 14 days. The intervention lotions were manufactured by mixing soil- and plant-based material (10%) to Aqualan L lotion. The soil- and plant-based material has been described in detail in our previous studies ^13,19,20,40^. The material contained composted ingredients comprising agricultural stack, gardening soils, deciduous leaf litter, peat, and *Sphangum* moss. The mixed and sieved material was saturated with ultra-pure mQwater and kept in the laminar for 4 h, covered with a lid but with holes in the side walls of the container to allow adequate air circulation. The soil–water mixture was then hand-squeezed using sterilized laboratory gloves over an ethanol-cleaned 250 μm sieve placed above another sterile plastic container. The extract was collected in separate 50 ml Falcon tubes, frozen at −20◦C overnight and freeze-dried for 48 h (Christ Alpha 1–4, B.Braun Biotech International). The participants were instructed to spread lotion with their dominant hand to the other arm from knuckles to the forearm and not to wash their hands for 15 minutes after that. Background information including medications, diseases, diets and living habits, were collected with questionnaires.

The study was conducted in accordance with the recommendations of the Finnish Advisory Board on Research Integrity, and an approval was received from the ethics committee of the Pirkanmaa Hospital District, Finland. In accordance with the Declaration of Helsinki, a written informed consent was obtained from all study participants. The trial has been registered in ClinicalTrials.gov (NCT03351543).

### Adverse events

Recorded adverse events included any undesirable effects, whether related or unrelated to the use of the lotions. Participants underwent regular dermatological examinations, and any changes in skin condition or the occurrence of adverse events were documented. Additionally, participants were encouraged to report any discomfort or changes they experienced during the trial.

### Skin sample collection and microbial analyses

Skin swabs were collected at baseline, after 14 days and after 35 days from the trial start to get follow-up data when lotion was not used. Skin swap samples were collected from dominant hand, i.e., from the same location of the forearm where the lotion was spread, with a sterile cotton-wool stick wetted in 0.1% Tween® 20 in 0.15 M NaCl (10 × 5 cm area, 10 seconds wiping). Samples were stored in −70°C until further processing.

DNA was extracted with DNeasy® PowerSoil® Pro Kit (Qiagen) according to the manufacturer’s standard protocol. PCR were performed for the V4 region within the 16S rRNA gene (three technical replicates from each sample) using 505F and 806R primers ^41^. Negative controls (sterile water) and a positive control (Cupriavidus necator JMP134, DSM 4058) were included in DNA extraction and PCR. Paired-end sequencing of the amplicons (2 X 300 bp) was performed on an Illumina Miseq instrument using a v3 reagent kit. Raw paired-end sequence files were processed into amplicon sequence variants (ASVs) using DADA2 ^42^ with the non-redundant Silva database version 138 ^43^. To conceptualize the different sequence read counts,samples were normalized with centered log-ratio (clr) transformation ^44^.

### Statistical analyses

All the statistical tests were done with R v4.3.1 ^45^. Linear mixed-effect models (LMM) [function lmer in lme4 package ^46^] were constructed to analyze temporal shifts in skin *Staphylococcus* taking into account pairing of intervention-placebo participants. To estimate differences in relative abundance of *Staphylococcus* between the treatments, we used interaction between treatment arms and time in the LMM model, as recommended ^47^. In detail, relative abundance of *Staphylococcus* was used as a dependent value, the interaction between treatment and time as an explanatory variable and paired participants as a grouping factor (Random factor) in LMM model.

### AD-trial: Experimental design

The study involved 12 adults, age ranging between 18-56 years. All of them had AD diagnosis and active eczema on the first day of the trial. Inclusion criteria were age between 18-65, legal competency and fulfillment of the Hanifin and Rajka’s criteria ^48^ for AD. Exclusion criteria were occupation in the agricultural sector, diagnosed cancer or ongoing cancer treatments, use of systemic immune suppressive medicines, phototherapy, solarium, or vacation abroad during the study or too severe eczema (pause not possible from AD medicines).

The study was conducted, and the data was collected in Helsinki, in facilities of Terkko Health hub, which is located at the Helsinki University Medical campus. Regional medical research ethics committee of HUS (Hospital District of Helsinki and Uusimaa) has given an affirmative statement for the study (diary number 15088/2022) on 28^th^ September 2022. Recruitment was started immediately. Written informed consent was provided by each volunteer before the participation. The trial has been registered in ClinicalTrials.gov (NCT06499766).

Restrictions during the study (28 days) were that on the designated test area it was not allowed to use other moisturizers or creams (including medical creams) than the test lotion. In the other parts of the body, it was allowed to use other creams including medical creams. AD medicines taken orally or by injection were prohibited during the study, as well as starting any new supplements that has not been part of the regular life before the study. A prewash period of 14 days was applied before the study, during which the use of any AD medicines was not allowed. On the clinical visit day, there was restrictions not to use alcohol or tobacco products 12h before the visit, and not to consume coffee, perform exercises causing sweating or wash the test area with soap 2h before the visit.

The study was randomized, double-blinded and placebo controlled. Each person had two test sites: one for the placebo and one for the inactivated microbial extract (Fig 1). Therefore, each volunteer served as both a control and a tester of the microbial extract. Randomization was conducted using a random number table by a person that was not involved to the study. She randomized the site of application of the lotions before the start, i.e. which lotion was used on which side of the body so that possible difference between the sites was not caused by dominating arm vs. non-dominating arm. The test lotions were made using Aqualan L as in pretrial and by mixing either plant and soil based inactivated material (later microbial extract), or coloring agent (MICA powder) to the lotion. The coloring ingredient were used to get the same brownish color to the placebo lotion as it was in microbial extract lotion. To maximize the safety, the proportion of the microbial extract, was 1% in AD-trial in contrast to 10% in pretrial. The microbial extract used in the AD-trial was manufactured by Uute Scientific Oy, Finland. The composition and diversity of the exposure materials used in both studies are corresponding and manufactured from similar raw materials. Volunteers and the research team were blinded until the results were analyzed.

**Figure 1.**
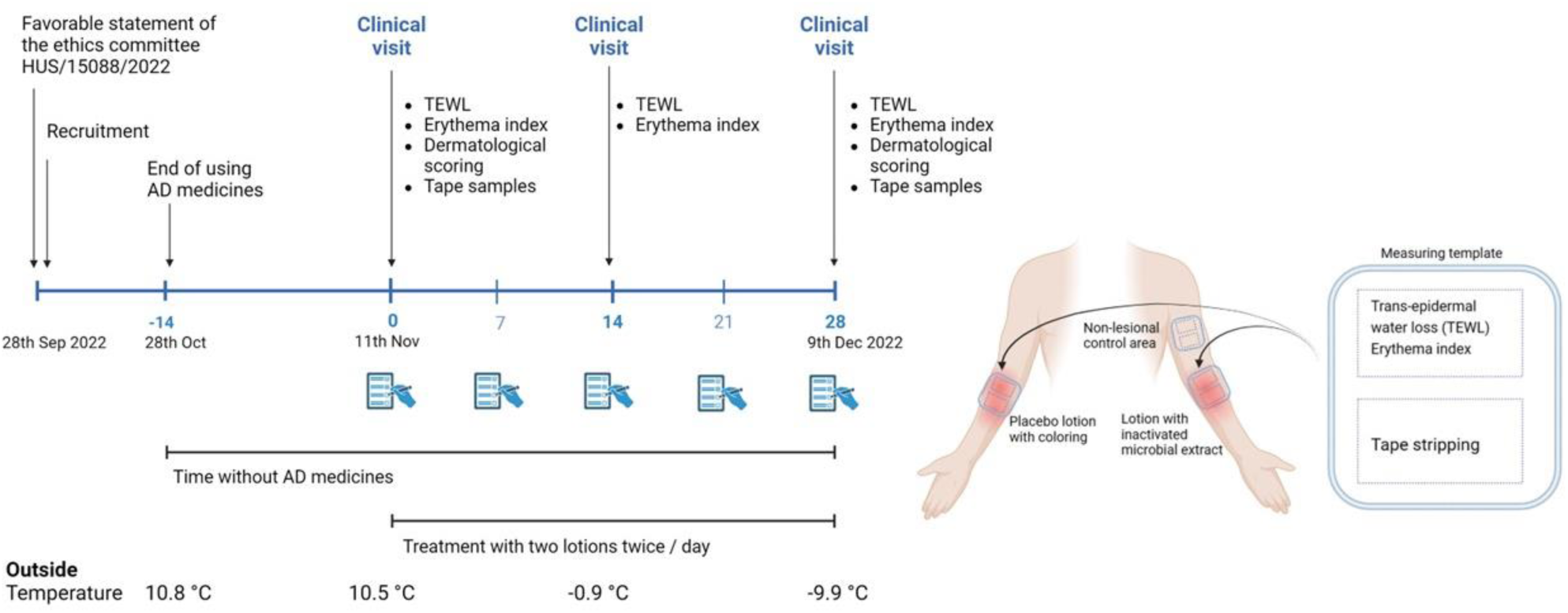
Trial protocol. Measurements were taken from both arms and on the non-lesional control area, on day 0, day 14 and day 28. Each participant had placebo lotion and lotion with inactivated microbial extract, which were used for selected arms daily during the 28-day trial. Mean outside temperature in Helsinki area is fetched from Finnish Meteorological Institute and presented to show the proceeding winter conditions. The image is created in BioRender.

Volunteers were met three times during the trial: on day 0, 14 and 28. The test lotions were used twice a day from day 1 to day 27 and only on the designated test areas. Analyses and sampling were done at the presented time points (Fig 1). TEWL was the primary outcome, and erythema index, dermatological scoring of atopic dermatitis and biochemical markers from tape strip samples were secondary outcomes. Outside temperature is presented because cold and dry weather, like in Northern Europe on winter times, increases the prevalence and risk of flares in AD patients ^49^.

### Dermatological evaluation and selection of test areas

At the start of the first clinical visit, volunteers met the dermatologist who confirmed the AD diagnosis and made the scoring of the eczema using two different methods. The eczema on whole body was scored using Atopic dermatitis severity index (SCORAD, 0-25 mild, 25-50 moderate, <50 severe) ^50^. This was done to follow the overall development of the eczema during the study, that reflects for example changes in the volunteers living environment (exposure to allergens, weather conditions etc.). Additionally, the atopic eczema was separately scored on both test sites by evaluating the six symptoms (redness, swelling, oozing/crusting, scratch marks, skin thickening, dryness) using the grade 0-3 for each symptom (0=clear, 1=almost clear, 2=moderate, 3=severe), maximum score being 18 per test site.

The test areas were chosen to be at active atopic eczema site and to be situated symmetrically on easily accessible locations on both body sides (primarily arms). Three measuring points were selected from the test areas: one lesional point from left and right side, and one non-lesional point from either of the site (Fig 1). Selected areas were photographed so that it was possible to take samples and measurements on the same points in the later meetings. Measurements were done first followed by tape stripping (Fig 1).

### TEWL and Erythema measurements and self-evaluation

Transepidermal water loss (TEWL, instrument Delfintech Vapometer) and Erythema index (Delfintech Skin Color Catch) were measured on the test and control sites. The TEWL measuring device uses closed chamber measurement principle. The sensor monitors the increase of relative humidity inside the chamber during the measurement phase and the evaporation rate value (g/m^2^h) is automatically calculated from the RH increase. ^51^. The result is mean of three repeated measures.

Participants fulfilled a self-evaluation questionnaire every seven days. The questionnaire included six questions of eczema symptoms and six questions about the user experience of the lotion (Supplementary Table 1). Participants were also interviewed in every meeting.

### Skin samples and biomarker analyses

We utilized a minimally invasive tape stripping suitable in detecting biomarkers ^52,53^. Skin tape strip sample was taken by placing 20 consecutive D-Squame tape strips (D100-D-Squame Standard Sampling Discs diameter 22 mm, Clinical and Derm LLC.) on selected spot and keeping each tape on place for 10 seconds gently pressing simultaneously with two fingers. The first two tapes were discarded and tapes 3.-20. were placed in the 1.5 ml test tubes (2 tapes per tube) with minimal overlapping. Tubes were put on dry ice until transferring them to the -80°C freezer.

Tape strip samples were thawed on ice and 800 µl phosphate-buffered saline (in house) containing 0.05 % Tween 20 (Sigma Aldrich) and protease inhibitor (Pierce™ Protease Inhibitor Mini Tablets, Thermo Fisher Scientific) was added to the first tube (containing tape strips 3. and 4.). The tube was mixed for 75 seconds using Vortex and the supernatant was then transferred to the next tube, which contained the tapes 5.-6. of the same sample series. The remaining tube with the tapes 3.-4. was spinned shortly to collect the residual supernatant. This procedure was repeated until the last tube of the same sample series was processed. Finally, the supernatant containing the combined samples from all the tapes 3.-20. was centrifuged 10 minutes at 4 °C using 16,100 RCF. The volume of this final sample was around 500 µl and it was divided into three aliquots stored at -80 °C for further analyses.

Cytokines were analyzed using Mesoscale U-plex® Biomarker Group 1 (Human) Multiplex assay (Meso Scale Diagnostics, LLC, USA). The measured cytokines were related to AD and they have been detected in skin tape strip samples from AD lesional skin in previous studies ^35,36,54^. The following cytokines were analyzed: IL-1β, IL-8, IL-13, TNFα, IL-18, IL-17E/IL-25, IL-22, IL-31, IL-33 and TSLP. The assay was performed according to the manufacturer’s instructions using MESO Quickplex SQ 120 instrument. Samples from the same person were analyzed in one assay to eliminate interassay variation. Results were analyzed with manufacturer’s program MSD Discovery Workbench. A blank tape sample was used as a control and samples were analyzed in two replicates. The concentration of cytokines was standardized for total protein concentration (Pierce™ BCA Protein Assay Kit; Thermo Fisher Scientific). Antimicrobial peptide Human Defensin Beta 2 was analyzed using Elabscience® Human DEFβ2 ELISA kit (Elabscience Biotechnology Inc., USA). Absorbance was read with Perkin Elmer VICTOR Nivo™ multimode plate reader.

### Statistical analyses

Statistical analyses were done using IBM SPSS Statistics and GraphPad Prism 9.0. The normality of the data was analyzed with Shapiro-Wilk normality test. Because number of cases was small and most of the variables were non-normally distributed, non-parametric tests were applied. Correlations between the different methods were analyzed using Spearman’s Rho. Comparison of fold changes between treated and placebo sites as well for difference of absolute values between the days 0 and 28 and between the sites (NL-Lesional sites) were done using Wilcoxon signed rank test.

## Results

### Pretrial

Safety profile was similar in live, inactivated and placebo treatment arms of the pretrial. There was no significant adverse effects associated with the use of live, autoclaved or placebo lotion in healthy volunteers. Both the intervention arm and the placebo group demonstrated comparable number of minor non-serious events, such as mild skin irritation, consistent with typical responses to skincare products. Participants in the live lotion arm reported a high level of tolerance as no severe adverse events or systemic reactions were observed throughout the trial.

There were no differences in the skin *Staphylococcus* abundance between pretrial treatment arms. Relative abundance of *Staphylococcus* on the skin stayed similar in all three treatment groups during the trial (Linear mixed model: p > 0.6; Table 2).

**Table 1.** Relative abundance of *Staphylococcus* on the skin in the three pretrial treatment arms.

| Treatment arm | Staphylococcus relative abundance |  |  |  |  |  |
| --- | --- | --- | --- | --- | --- | --- |
|  | Day 0 |  | Day 14 |  | Day 35 |  |
|  | Mean | sd | Mean | sd | Mean | sd |
| Live lotion | 1034 | 36 | 1026 | 22 | 1017 | 22 |
| Autoclaved lotion | 1015 | 17 | 1019 | 19 | 1018 | 19 |
| Placebo lotion | 1013 | 12 | 1010 | 8 | 1009 | 9 |

**Table 2.** Linear mixed model (LMM) was constructed to analyze the change difference between treatment arms A) between day 0 and 14 and B) between day 0 and day 35. LMM statistics are reported as estimates for fixed effects, standard error (se), t value and probability p value.

| A) LMM day 14 | Estimate | se | t value | p value |
| --- | --- | --- | --- | --- |
| Autoclaved vs placebo | 7.49 | 15.61 | 0.48 | 0.63 |
| Live vs placebo | -4.50 | 16.64 | -0.27 | 0.79 |
| B) LMM day 35 | Estimate | se | t value | p value |
| Autoclaved vs placebo | 7.60 | 15.61 | 0.49 | 0.62 |
| Live vs placebo | -13.12 | 16.64 | -0.79 | 0.43 |

### Atopic Dermatitis trial

All 12 volunteers were randomly assigned to the study, 11 received the intended treatments and 10 came to all the intended visits. One volunteer started to use a corticosteroid creme on the research areas during the study so primary outcome (TEWL) and secondary outcomes are from 9 volunteers that didn’t use any other creams or lotions than test lotions on research areas. Subjective evaluation includes results from all 11 volunteers that received the treatments.

### Self-evaluation

In the interviews carried out on days 14 and 28 after the initiation of the treatment, the participants reported their skin becoming drier needing more moisturizing when winter started. They also reported that the test lotion alone (Orion Aqualan L + either containing exposure material or control) was not moisturizing enough. Overall, participants named cold weather (winter was progressing during the study; Fig 1), common cold, stress, alcohol, swimming, cleaning chemicals and renovation dust as exacerbating factors during the trial.

Participants reported temporary itching, redness and feeling of heat right after lotion application on the first days of use as follows: on both test sites n=5, only at treated site n=2, only at placebo site n=1. On day 14 and day 28 visits only one person still had temporary itching existing only a short period of time right after the lotion application and only at treated site. No other skin related treatment-associated adverse effects were reported. Three persons reported that the brown color of the lotion was staining textiles. According to the questionnaires, there were no significant differences between the user experience or skin condition between the microbial extract and placebo treated skin areas.

### Scoring of the eczema

Participants’ atopic eczema was mild (SCORAD 0-25) or moderate (SCORAD 25-50) before and after the trial (Fig 2A). SCORAD values indicating the whole-body area eczema showed a nonsignificant decreasing trend during the trial ranging from baseline median level of 24 to 17 at day 28. The local scoring value of tested skin area did not change as median score was 2 in both sites at days 0 and 28, respectively (Fig 2A).

**Figure 2.**
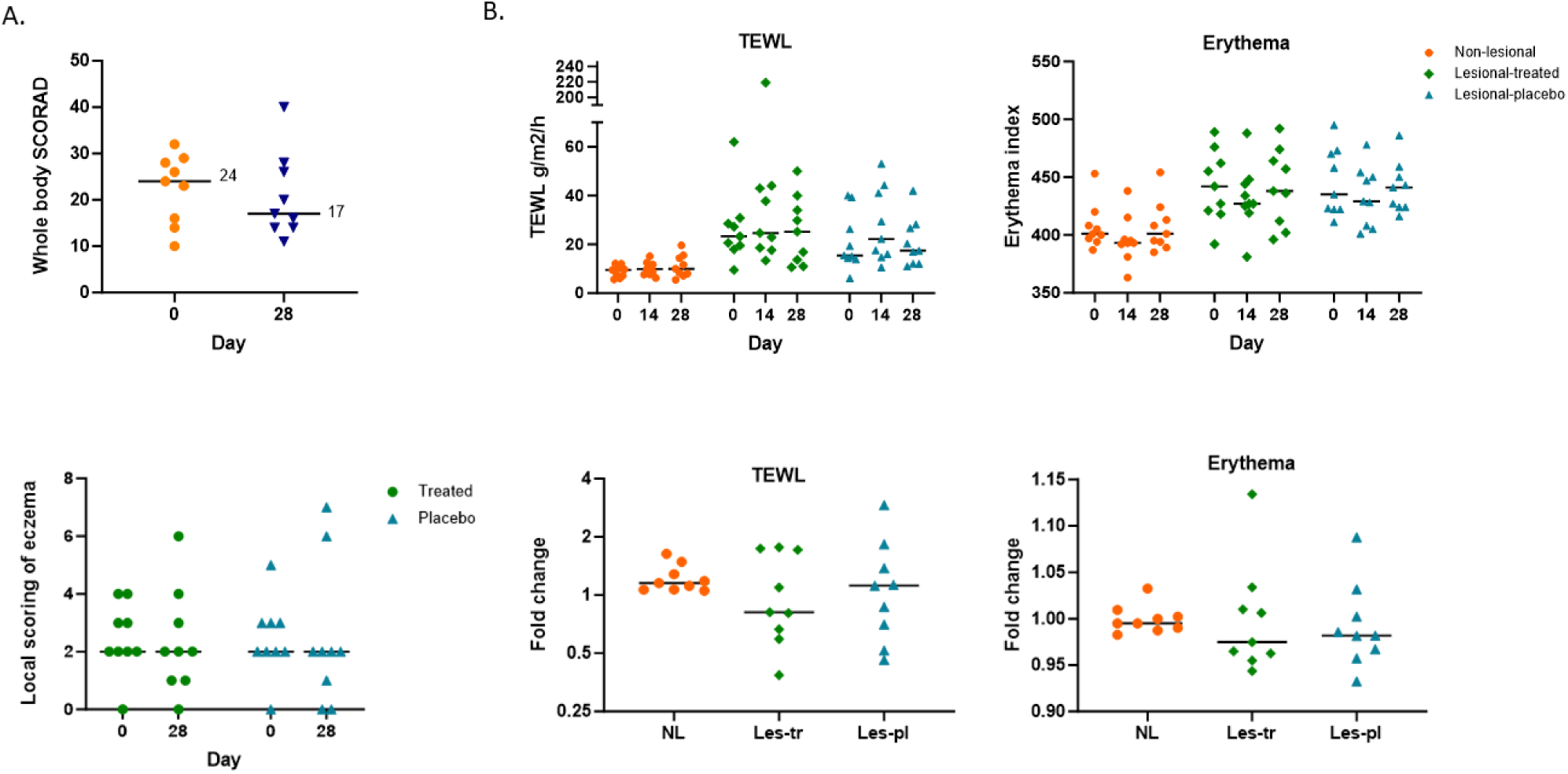
**A. Whole body SCORAD points before and after the intervention trial of atopic eczema on whole body and local scoring of eczema separately on treated and placebo skin sites before and after the biodiversity intervention.** Scoring was done by evaluating the appearance of the six symptoms (redness, swelling, oozing/crusting, scratch marks, skin thickening, dryness) using the grade 0-3 for each symptom (0=clear, 1=almost clear, 2=moderate, 3=severe). **2.B. TEWL ja Erythema index illustrated as absolute values and fold changes (end value/start value).** NL= non-lesional site, Les-tr = Treated lesional site, Les-pl = placebo lesional site.

### Transepidermal water loss and Erythema index

TEWL was studied in all three defined body areas (Fig 2B). Non-lesional skin TEWL values were significantly lower compared to lesional skin site on both test sites (p-values 0.008 - 0.015) indicating reliability of the method. The erythema index was measured from the same sites as the TEWL. Also, Erythema values were significantly lower on non-lesional skin compared to lesional skin test sites (p-values 0.008 - 0.011). When only lesional sites were analyzed, there were no statistically significant differences between the two lesional test sites nor inside each lesional test site during the trial (Fig 2B). Fold change for TEWL and Erythema is calculated by dividing the end value by the start value (Fig 2B). On the lesional test areas there were lot of variation in TEWL fold change values, but median level on the site treated with the extract containing lotion was slightly decreased. Erythema levels did not change on non-lesional site and median levels slightly decreased on both lesional sites during the trial (Fig 2B).

### Biochemical markers

There was a statistically significant decrease in the concentration of IL-1β (p=0.028) and hBD-2 (p=0.008) during the trial on placebo site (Fig. 3A and 3B). When cytokine concentrations were compared between lesional and non-lesional skin areas (n=18) they were significantly higher on lesional sites before and after the trial (Supplementary Table 2). There were no significant differences in cytokine or hBD-2 fold changes between treatment sites during the study (Fig 1A and 1B in the Supplementary material).

**Figure 3.**
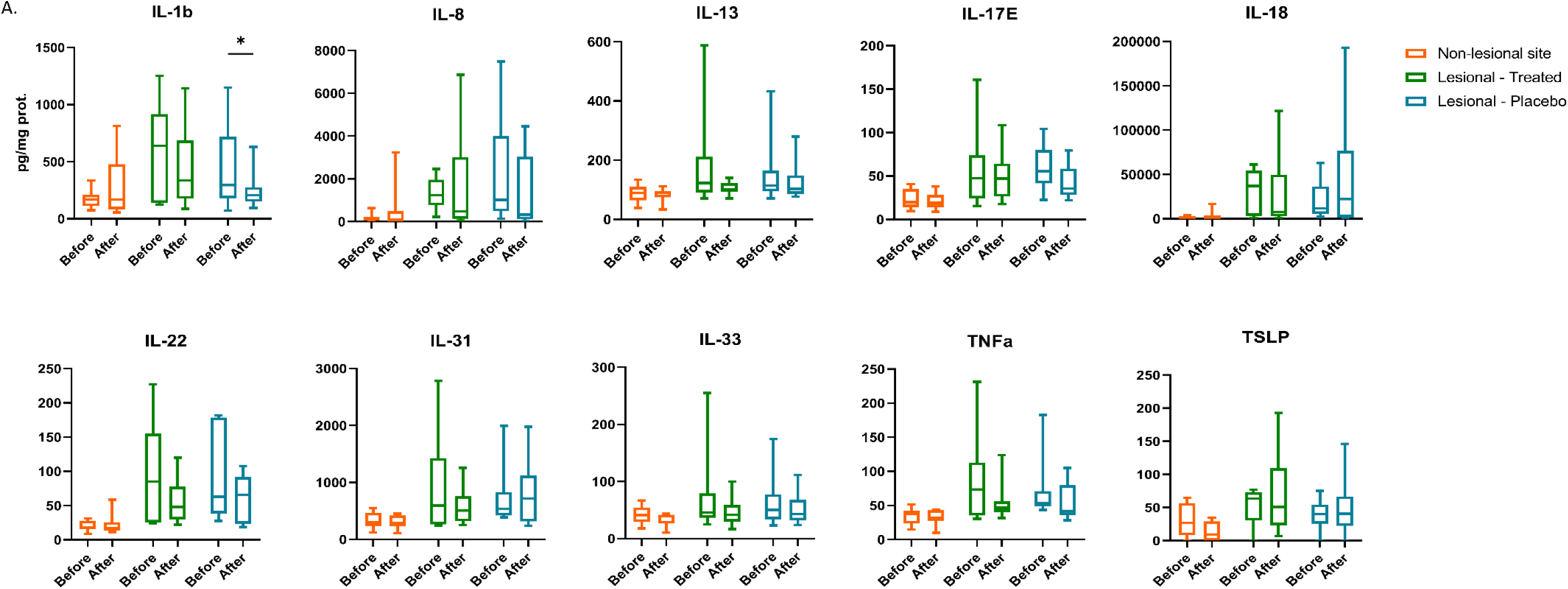
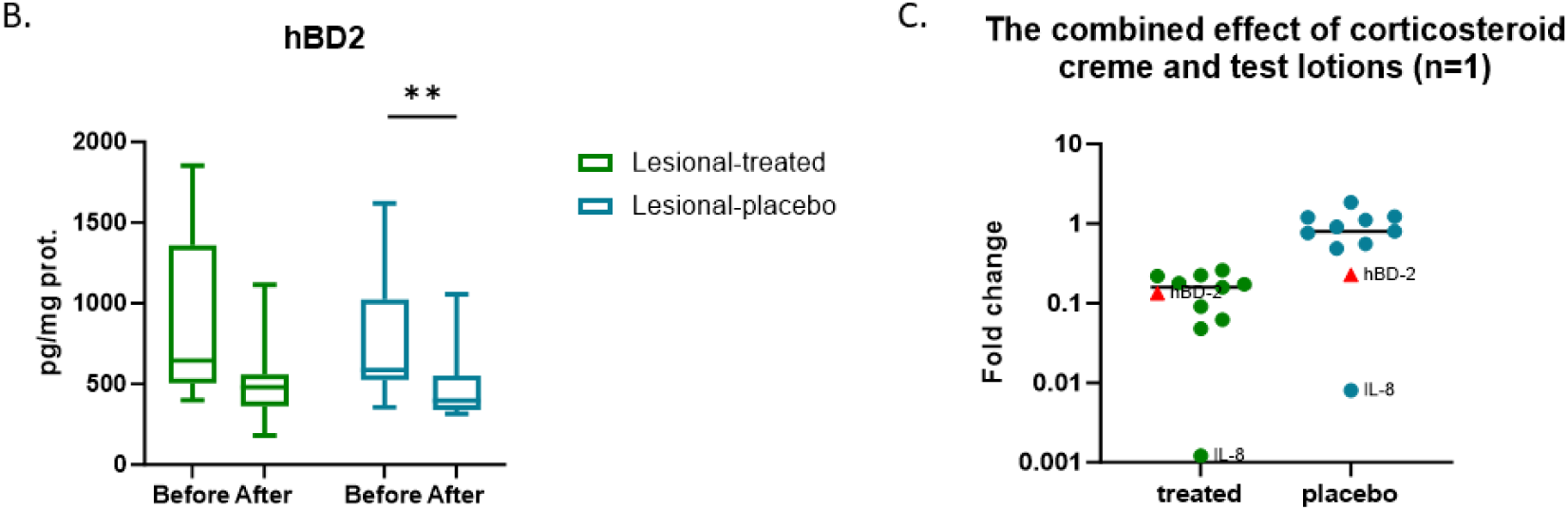
**A**. Concentration of ten pro-inflammatory cytokines during the trial in tape strip samples (n=9) from non-lesional and lesional skin sites. **B.** Concentration of human β-defensin during the trial in tape strip samples (n=9) from lesional skin sites. **C.** Fold change of cytokines (dots) and human β-defensin 2 (red triangles) when corticosteroid crème was taken into use parallel with test lotions by one person in the middle of the trial.

One of the participants, that had the highest SCORAD value at Day 0 (SCORAD 36), started to use Bemetson K (corticosteroid crème with betamethasone and clioquinol) at Day 16 in addition to the test lotions and their samples were analyzed separately (Fig 3C).

### Both test sites

When the lesional test sites (placebo and treated) were analyzed together (n=18), there were statistically significant decrease of IL-1β (p=0.01), IL-22 (p=0.043), IL-33 (p=0.037) and hBD2 (p=0.002) between days 0 and 28 (Fig 4), and a decreasing trend of IL-13 (p=0.059) and TNFα (p=0.06). The lotion itself (placebo) and the microbial extract alleviated together Th2, Th17 and Th22 type allergic responses (Fig 4).

**Figure 4.**
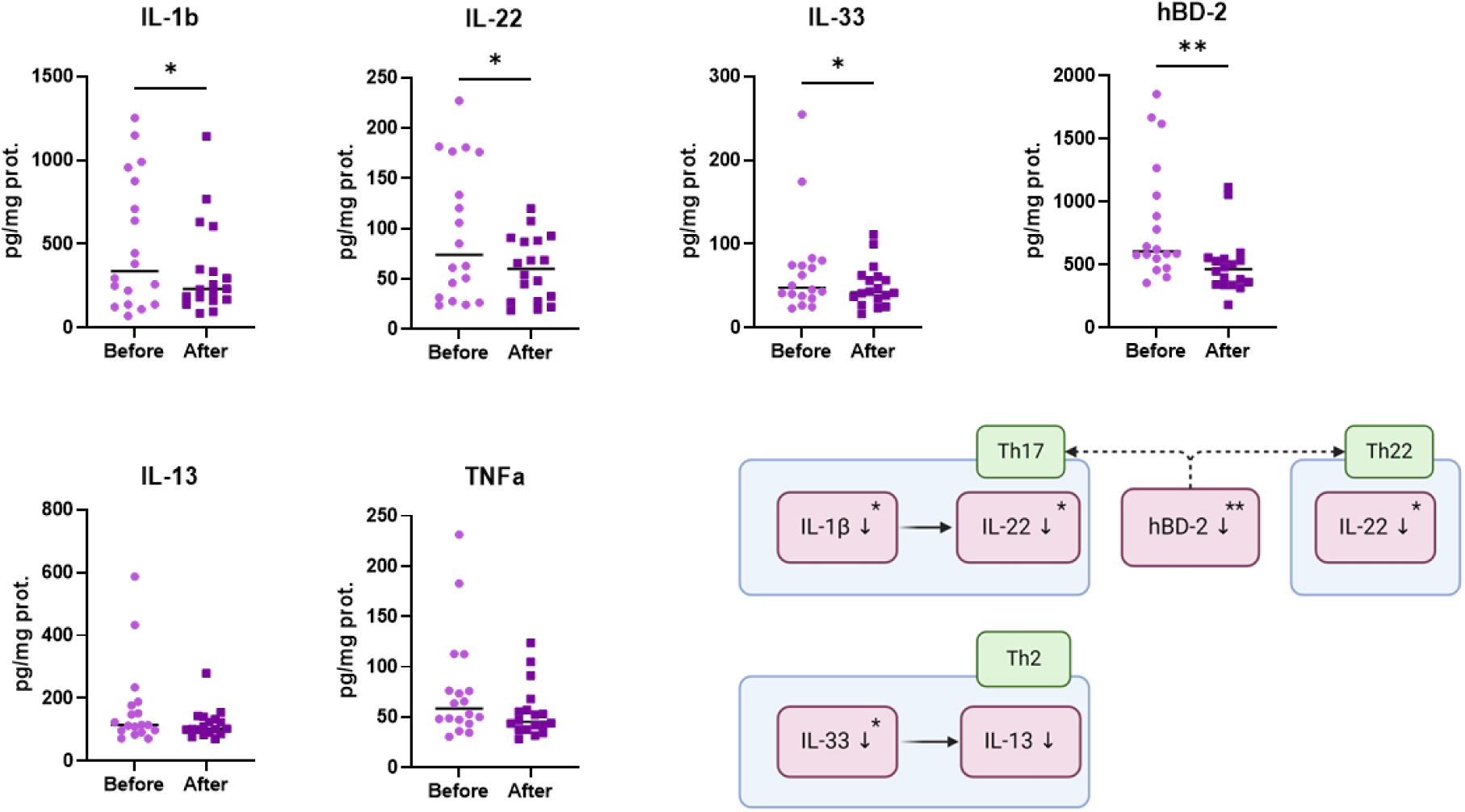
Both test sites combined. IL-1β (p=0.01), IL-22 (p=0.043), IL-33 (p=0.037), hBD2 (p=0.002), IL-13 (p=0.059), TNFα (p=0.06). Relation of the analyzed biomarkers to T helper (Th) cells and the role of hBD-2 as a chemoattractant for Th17 and Th22 cells. Part of the image is created in BioRender.

### Correlations

Many of the analyzed biomarkers (cytokines, hBD-2) and physical measurements (TEWL, Erythema index) correlated significantly with each other (Fig 5). Correlation was analyzed using results from both lesional sites and days (0 and 28) in the analysis (n=40).

**Figure 5.**
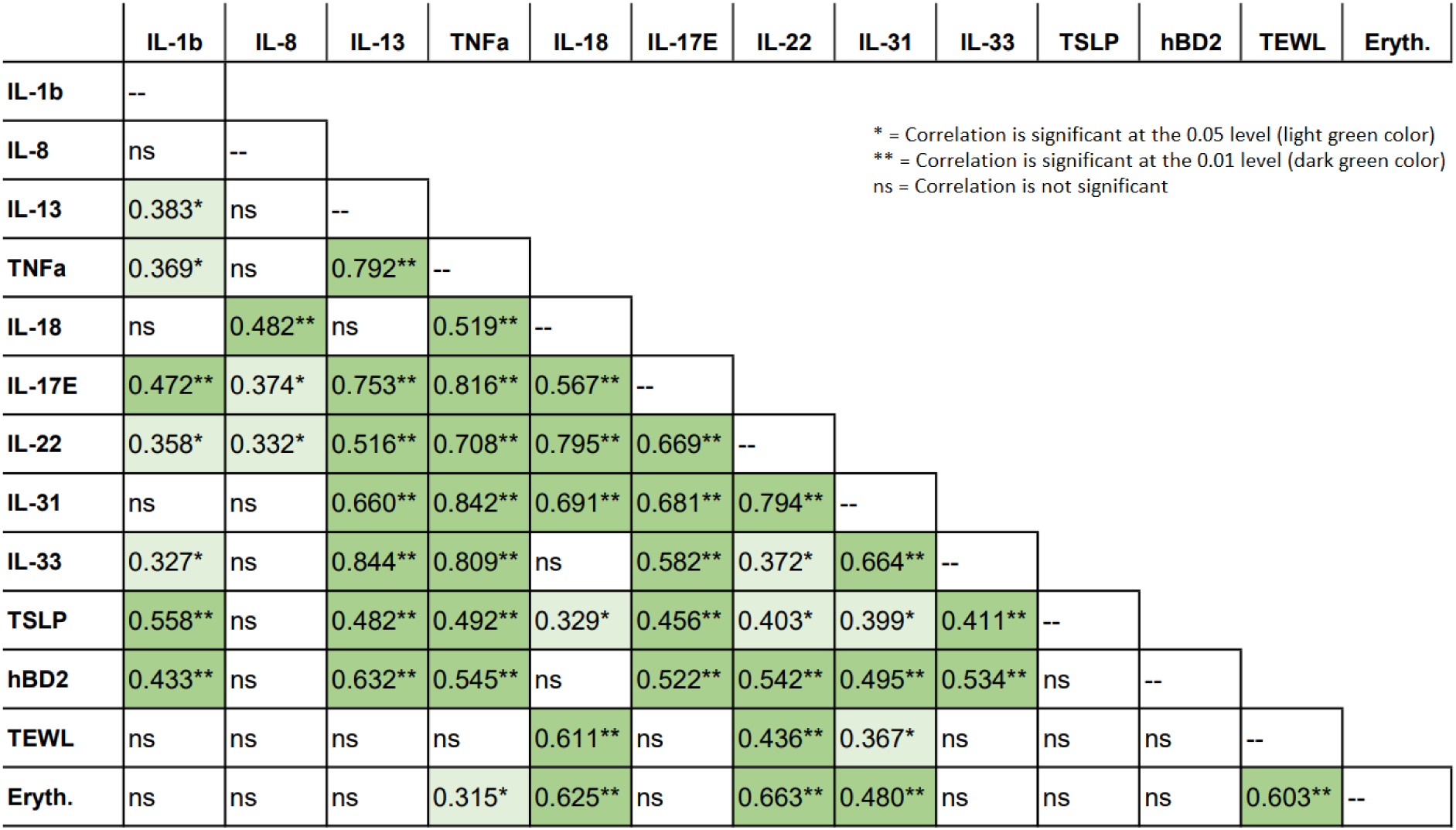
Spearman’s rho Correlation Coefficients (rs) between the different methods or analytes with lesional skin samples (n=40 including before and after values on both lesional test sites). * Correlation is significant at the 0.05 level (2-tailed). ** Correlation is significant at the 0.01 level (2-tailed). ns= correlation is not significant.

## Discussion

As there is a clear need for new nonprescriptive treatment options for AD, we wanted to study whether nature exposure can be a supportive option to other treatments. The impetus behind this idea derives from biodiversity hypothesis and the fact that exposure to highly diverse microbes in childhood protects from atopy and allergies ^18,27–29^. Our previous studies have shown positive immunological outcomes when contacts to natural highly diverse microbes have been recovered later in life ^13,19^. The main research question was to find out, if microbial extract in moisturizer is safe and feasible way to introduce natural microbes to atopic skin. This was approached in a two-phase study: first a trial with healthy adults with a high dosage using live and inactivated microbes (pretrial), and after this exposure was found to be feasible with no adverse effects, we proceeded to AD patients with lower dosage and using only inactivated microbial extract (AD-trial). The secondary aim was to study the possible immunological effects of the exposure.

The pretrial demonstrated that the use of lotion for two weeks with live microbes or autoclaved microbial extract had a favorable safety profile with no significant adverse effects and no change in *Staphylococcus* relative abundance. It was also proved to be a feasible to administer microbial extracts as a skincare product. The AD-trial supported this conclusion, as there were no significant differences in the profiles of reported adverse effects, local scoring of atopic eczema, TEWL or erythema levels between the placebo and the microbial extract of the participants’ research areas.

In AD-trial we found no significant reduction of pro-inflammatory cytokines on the treated skin site alone. However, there was a lot of variances in the fold change results on the treated site especially with cytokines IL-13, IL-22, IL-31, IL-33 and TNFα compared to placebo and non-lesional skin sites. This may indicate that some people respond more favorably to lotion with microbial extract. It has been demonstrated that genotype affects the host response on environmental and LPS exposure ^27^ potentially explaining this observation. Based on the previous studies, systemic immunological effects could be expected after biodiversity intervention ^12,13,55^. The weakness of the AD-trial protocol to study systemic effects is that there was no independent control group, but the control sites were in the same persons as intervention sites.

In AD-trial, systemic immunological responses based on exposure and/or increased use of moisturizers were noticed. The simultaneous downregulation of IL-1β, IL-22 and hBD-2 expression is logical, as they all affect the Th17 cells (Fig 4). Th17 cells control immune responses against extracellular bacteria and fungi ^56^. IL-1β promotes differentiation of naïve T-cells into Th17 cells that in turn produce IL-22 ^57^. IL-22 is also connected to an independent subset of Th22 cells, and it triggers hBD-2 antimicrobial peptide release from keratinocytes ^58,59^. hBD-2 controls activity against bacteria, fungi and viruses, as well as regulate immune responses ^60^. IL-22 has been demonstrated to downregulate the production of many skin barrier proteins like keratin, loricrin and profilaggrin, which are needed to recovery of the epidermis ^61^. Another previous study showed that the expression of IL-22 is elevated in the inflamed skin in AD patients ^62^. Interestingly, we showed the reduction of IL-22 on lesional skin sites (Fig 4) further supported by its correlation between TEWL and erythema indexes (Fig 5).

A common factor for Th17 and Th22 cell subsets is a chemokine (C-C motif) receptor 6 (CCR6) expressed on these cells. The ligand for CCR6 is chemokine (C-C motif) ligand 20 (CCL20) ^58^, and the expression of both CCL-20 and CCR6 are upregulated in AD ^63^. It has been demonstrated *in vitro* that scratch injury to epidermal keratinocytes upregulates the production and release of CCL20, which chemoattracts Th17 immune cells ^64^. On the contrary, the lowered concentrations of Th17 and Th22 related cytokines and hBD2, which also recruits Th17 and Th22 cells, during the AD-trial indicated the potential to induce tolerance and mitigate the AD symptoms. Constant activation of epithelial cell TLR receptors may create immunological tolerance which could be seen here as Th17 cells and hBD-2 both set immune responses against extracellular bacteria and fungi. It remains to be further demonstrated if the reduction of IL-1b, IL-22 and hBD-2 reflects reduction of CCR6 expressing Th17 and Th22 cells during biodiversity intervention or if it is an indication of increased use of skin moisturizers.

Another finding of the present study was lowered concentration of Th2 associated IL-33 on treated and placebo sites. IL-33 is also connected to AD ^65,66^. IL-33 activates Th2 cells that produce IL-13 ^57,66^, which is in line to our result that there was strong correlation between IL-33 and IL-13 on lesional skin (rs=0.884, Fig 5). IL-33 is also linked to pruritus and decreased skin barrier function. Therefore, its reduction during the study is an additional indication of the mitigation of atopic eczema due to microbial exposure and/or better moisturization. AMPs, especially hBDs, can also induce Th2 cells to express IL-31, which is another pruritogen and thereby reduction of hBD-2 level during the trial is a good thing for atopic dermatitis patients ^60^.

Interestingly, one of the participants of AD-trial used Bemetson K (corticosteroid) during the last 12 days of the trial, and we found a strong decrease in participants’ cytokine levels between day 0 and day 28 as well as hBD-2 levels in the treated site as compared to placebo site. These results indicate that the reduction of hBD-2 is related to mitigation of atopic eczema and related inflammation. Although this was a result of only one participant, it is intriguing to speculate if the nature exposure could function synergistically with corticosteroids and provide in the future a complementary treatment modality with the corticosteroids thus lowering the required dose and thereby reducing the adverse effects of long-term corticosteroid use.

Cytokine IL-8 had the weakest correlation to other cytokines, which is in line with the previous findings where Th2 cytokines, like IL-13, have been shown to suppress IL-8 production ^67,68^. The correlation of TEWL, indicating the skin barrier function, with cytokines IL-18, IL-22 and IL-31, has been previously shown ^54^, and furthermore, IL-31 disrupts skin barrier formation by downregulating filaggrin expression ^69,70^ and increasing itch ^60^.

Selecting the right vehicle is critical in testing ingredients in these kinds of trials. As participants reported, the used lotion was too light, meaning that on winter, a cream that contains more oil, should have been a better choice as the only moisturizer. Considering the fact that we used relatively light lotion, and the winter was proceeding during the AD-trial typically worsening the symptoms of AD, it was noteworthy that SCORAD values still decreased during the trial. It is possible that when participants committed to the study and used moisturizing test lotions regularly, the observed effect on both sites (four biochemical markers decreased significantly) was because the skin was more properly moisturized, which consequently mitigated inflammation and atopic eczema.

As a conclusion, we found that inactivated microbial extract in lotion is safe and feasible way to administer biodiversity exposure to healthy participants and AD patients. Notably, we found that antimicrobial peptide hBD2 was not increased by the intervention, rather it decreased together with several pro-inflammatory cytokines associated to AD. Weakness of the AD-trial was that we could not separate the possible systemic effect of the biodiversity intervention from enhanced use of moisturizer because there was no separate control group. Simultaneous use of corticosteroid creme and the lotion containing microbial extract led to stronger decrease of the pro-inflammatory cytokines and hBD-2 compared to using only the lotion with corticosteroid, but this needs to be further tested as it is based on the data from only one person. The results of the present study contribute valuable information to support the safety and feasibility profiles of biodiversity exposure in AD patients. They also establish a strong foundation for the use of biodiversity intervention in forthcoming trials.

## Data Availability

All data produced in the present study are available upon reasonable request to the authors

## Acknowledgements

We thank PhD Niila Jouppila from Tampere University and scientists of MesoScale Diagnostics for guidance to multiplex assay analysis. A special thank is owed to the study participants for their participation in this study. The authors wish to acknowledge CSC – IT Center for Science, Finland, for computational resources, and FIMM Genomics NGS Sequencing unit at University of Helsinki supported by HiLIFE and Biocenter Finland.

## Author contributions

J.K. participated in the planning and implementation of the AD trial, enrolled and assigned the participants to the study, performed the analyzes, prepared the figures and tables, and wrote the manuscript. I.M. participated in the planning, implementation and analysis of the result of the AD trial.

L.K. participated in the planning and analysis of the AD trial results. M.R. participated in the planning, implementation and analysis of the pretrial and manuscript preparation. H.Hu. participated in the analysis of the AD trial results. R.P. participated in the planning and implementation of the pretrial and manuscript preparation. P.A. participated in the implementation of the AD trial. H.Hy. participated in the planning and analysis of the AD trial results. V.H. participated in the analysis of the AD trial results and manuscript preparation. A.S. participated in the planning and analysis of the trials. O.H.L. participated to the planning and the conduction of the trials, analysis of the results and manuscript preparation.

## Competing interests

The study was conducted at the Doctoral School of Industry Innovation (DSII) at Tampere University in collaboration with and partially funded by company Uute Scientific Ltd, Helsinki, Finland.

A.S., H.Hy., O.H.L., J.K. and I.M. are co-owners of Uute Scientific, which produces microbial extract.

