## Supplemental data for "Testing the Safety of the Nature Based Microbial Exposure with Atopic Dermatitis Patients – A Randomized, Placebo Controlled, Double-Blinded Pilot Trial"

**Table 1. Self-evaluation questionnaires** (filled every 7 days during the trial)

Please choose an option that best describes the situation on test area during the last week.

1 = strongly disagree, 2 = disagree, 3 = agree, 4 = strongly agree

| **A. Skin condition on test area during the last week** | **1** | **2** | **3** | **4** |
|---|---|---|---|---|
| 1. Skin has been itchy. | | | | |
| 2. Skin has felt dry. | | | | |
| 3. Skin has been flaky. | | | | |
| 4. There has been irritation (redness, feel of heat, itching) on skin. | | | | |
| 5. There has been ulceration and cracking on skin. | | | | |
| 6. My sleep has been disturbed because of the skin. | | | | |
| **B. Experience of the lotion on the test area during the last week** | **1** | **2** | **3** | **4** |
| 7. The test lotion has been pleasant to use. | | | | |
| 8. I have not missed other creams. | | | | |
| 9. I have not missed medical creams. | | | | |
| 10. Two times a day has been enough for moisturizing the skin. | | | | |
| 11. The lotion has been helpful for my eczema. | | | | |
| 12. Skin reactivity to external factors (like allergens and washing) has been decreased. | | | | |

Supplementary material for the manuscript:

Testing the Safety of the Nature Based Microbial Exposure with Atopic Dermatitis Patients – A Randomized, Placebo Controlled, Double-Blinded Pilot Trial

**Table 2.** Comparison of the difference between non-lesional (NL) and lesional (Les) skin tape strip samples in concentration of pro-inflammatory cytokines. Comparison is done between non-lesional and lesional (n=18) and between non-lesional and treated (n=9) and non-lesional and placebo (n=9). Statistical test used is Wilcoxon Signed Rank test for two related samples.

* Difference is significant at 0.05 level
** Difference is significant at 0.01 level
*** Difference is significant at 0.001 level
ns = not significant

| | NL vs. Lesional (n=18) | | NL vs. Les.treated (n=9) | | NL vs. Les.placebo (n=9) | |
|---|---|---|---|---|---|---|
| | day 0 | day 28 | day 0 | day 28 | day 0 | day 28 |
| IL-1b | ** | ns | * | ns | * | ns |
| IL-8 | *** | *** | ** | ** | ** | * |
| IL-13 | *** | ** | * | * | ** | ns |
| TNFa | *** | ** | * | * | ** | ns |
| IL-18 | *** | *** | ** | * | ** | * |
| IL-25 | *** | *** | * | * | ** | * |
| IL-22 | *** | *** | ** | ** | ** | * |
| IL-31 | *** | *** | * | ** | ** | * |
| IL-33 | * | * | ns | ns | ns | ns |
| TSLP | * | ** | ns | ** | ns | * |

Supplementary material for the manuscript:

Testing the Safety of the Nature Based Microbial Exposure with Atopic Dermatitis Patients –
A Randomized, Placebo Controlled, Double-Blinded Pilot Trial

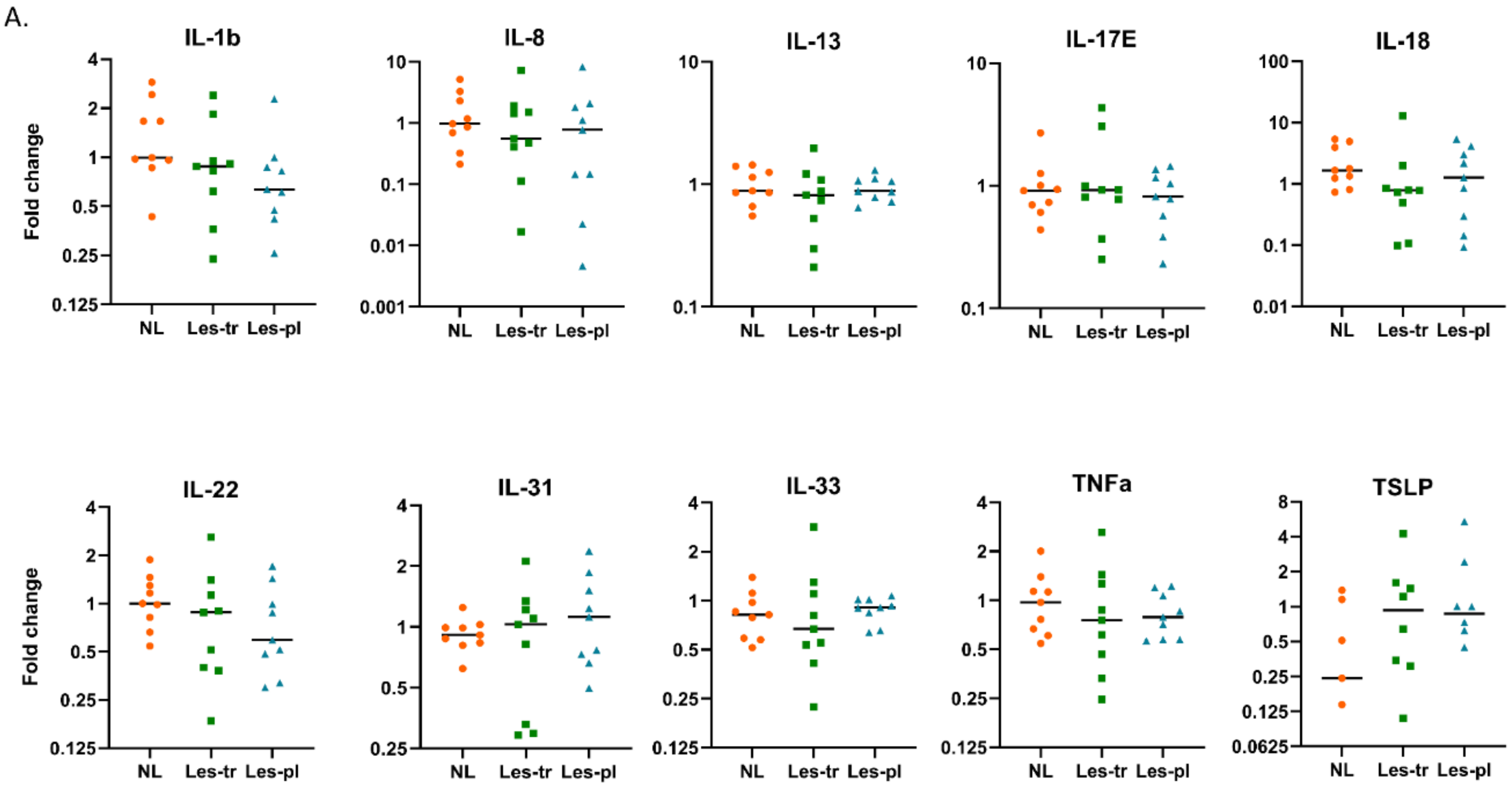

Supplementary material for the manuscript:

Testing the Safety of the Nature Based Microbial Exposure with Atopic Dermatitis Patients – A Randomized, Placebo Controlled, Double-Blinded Pilot Trial

B.

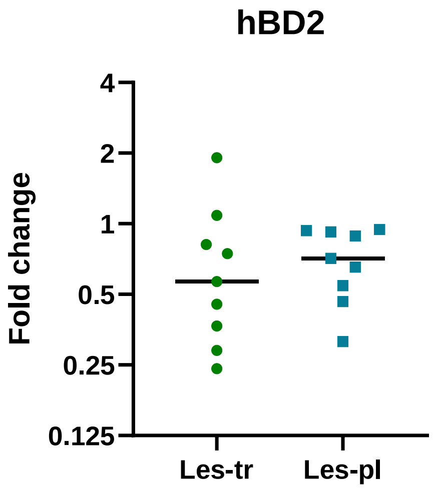

**Figure 1. A. Fold changes (end value/start value) during the trial. A.** Fold change of pro-inflammatory cytokines in non-lesional (NL) skin, lesional treated site (Les-tr) and lesional placebo site (Les-pl). **B.** Fold change of human β-defensin 2.
